# Complexity of baseline cognitive and psychological status in haematology patients planned for chimeric antigen receptor T-cell therapy

**DOI:** 10.1101/2023.06.03.23290918

**Authors:** Valeriya Kuznetsova, Hannah Rosenfeld, Carmela Sales, Samantha van der Linde, Izanne Roos, Stefanie Roberts, Fiore D’Aprano, Samantha M Loi, Mark Dowling, Michael Dickinson, Tomas Kalincik, Simon J Harrison, Mary Ann Anderson, Charles B Malpas

## Abstract

**Background:** Immune effector cell-associated neurotoxicity syndrome (ICANS) is a relatively common consequence of chimeric antigen receptor T-cell (CAR-T) therapy, with a wide range of possible cognitive presentations. The aim of this study was to characterise a real-word cognitive and psychological status of patients with advanced haematologic and solid organ malignancies planned for CAR-T. We also aimed to examine utility of two cognitive screening approaches.

**Methods:** Patients underwent specialist cognitive assessment, including a self-report questionnaire of psychopathology and subjective cognitive function. A subset of individuals also completed the Montreal Cognitive Assessment (MoCA).

**Results:** Of 60 patients included, 15-16 (25%-27%) presented with evidence of cognitive impairment, with six unique patterns of dysfunction. Impaired patients were more likely to have B-cell acute lymphoblastic leukaemia (*BF_10_*=9.30), be younger (*BF_10_*=7.76), have bone marrow involvement (*BF_10_*=5.18), report history of anxiety (*BF_10_*=4.85), or have evidence of psychopathology (*BF_10_*=31.30). Analyses did not support the utility of cognitive screening. Of those patients who completed a self-report measure of psychopathology, nine (15.8%) were elevated on at least one symptom domain.

**Conclusions:** The findings demonstrate a broad spectrum of dysfunction and psychopathology in this cohort, emphasising the importance of baseline evaluation for detecting cognitive neurotoxicity symptoms that might arise after CAR-T infusion.

## Introduction

The immune effector cell-associated neurotoxicity syndrome (ICANS) is a common and potentially life-threatening side-effect of chimeric antigen receptor T-cell (CAR-T) therapy, which is now a standard of care for some relapsed or refractory haematological malignancies.^1, 2^ The syndrome has a diverse range of possible neurological and cognitive sequalae, and current detection and classification approaches lack diagnostic sensitivity and specificity.^2, 3^ Cognitive status is not well characterised in patients prior to CAR-T infusion, adding a layer of complexity to distinguishing new symptoms from possible pre-existing dysfunction.

Both disease itself and cancer treatments are commonly associated with cognitive impairment.^4^ Dysfunction can vary, spanning processing speed, attention, learning and memory, executive, and language functions.^5–7^ Patients become eligible for commercial CAR-T therapy if their cancer is refractory to or has progressed following prior lines of systemic therapy or stem cell transplantation. All patients have been exposed to one or more prior chemotherapy regimens, with many involving central nervous system (CNS) penetrating agents. Typical treatment history in this cohort suggests that some patients could have pre-existing cognitive dysfunction. Psychopathology (e.g., depression or anxiety) can also contribute to transient disruption in cognition and is prevalent in cancer population.^8–10^ Thus, evaluating cognitive and psychological status prior to receiving CAR-T therapy is necessary to adequately distinguish neurocognitive consequences of ICANS from alternative aetiologies.

Hospital resources can be limited, and a baseline specialist cognitive assessment might not be possible for every patient. A screening approach could be used to identify individuals with suspected dysfunction to be referred for further examination. The Montreal Cognitive Assessment (MoCA) is commonly used as a cognitive screen in many clinical situations, including in patients with haematological cancers.^6, 11–13^ Subjective cognitive concerns are common in the cancer population and could serve as another way of detecting objective impairment.^14, 15^ Diagnostic utility of perceived cognition was demonstrated by Buckley et al.,^16^ who found that phenomenology of a subjective complaint could distinguish individuals with mild cognitive impairment from healthy controls. Thus, both the MoCA and a measure of subjective cognitive function are reasonable candidates for cognitive screening prior to CAR-T.

The current study aimed to characterise cognitive and psychological status in CAR-T patients pre-infusion. A substantial proportion of these individuals are expected to present with evidence of objective cognitive dysfunction and psychopathology. We also aimed to evaluate utility of the MoCA and a self-report psychometric measure of subjective cognitive function in detecting impairment. Capturing a comprehensive baseline pre-CAR-T will aid future investigation of clinical manifestation of ICANS and improve neurotoxicity management and outcome prognostication.

## Method

### Participants

Prospective cohort study of patients who were scheduled for CAR-T therapy at Peter MacCallum Cancer Centre haematology service between May 2021 and January 2023. The treatment was either planned as a commercial product for eligible patients or as part of a clinical trial. All patients underwent a routine specialist cognitive assessment prior to the infusion. Findings from these assessments were accessed, including the nature of cognitive complaints and psychometric profiles. Demographic and clinical characteristics were obtained from patient charts. Patients were included if they were at least 18 years of age, were proficient in English, and did not have an intellectual disability or a history of significant neurological illness. Approval was granted by the Peter MacCallum Cancer Centre Human Research Ethics Committee (21/145). A request for waiver of the requirement for consent was approved for this cohort. The study conforms with World Medical Association Declaration of Helsinki.

### Cognitive assessment

The cognitive assessment comprised a clinical interview to elicit a cognitive complaint and psychometric examination. A subset of patients completed a self-report questionnaire of psychopathology and subjective cognitive function (SPECTRA Indices of Psychopathology).^17^ A screening measure of objective cognitive impairment (MoCA) was also administered to a subset of patients.

### Psychometric examination

A battery of psychometric cognitive instruments was selected to capture a wide range of possible dysfunction. Five cognitive domains (i.e., processing speed and attention, executive function, memory, language, and visuospatial function) were examined in accordance with recommendations from the International Cognition and Cancer Task Force (ICCTF).^18^ Raw scores were converted into standardized z-scores based on the demographically adjusted normative data. A test score was classified as impaired if the z-score was <= −1.5 standard deviations.^18, 19^ A domain was classified as impaired if at least two scores were impaired (or one score for the visuospatial function domain) or a z-score averaged <= −1.5 across tests in that domain. A patient was classified as cognitively impaired if at least one domain was impaired. Table 1 in the supplementary materials describes all the instruments.

**Table 1.** Baseline clinical characteristics

| Characteristic | *M(SD)* | *Range* | Characteristic | *M(SD)* | *Range* |
| --- | --- | --- | --- | --- | --- |
| Age (years) | 57.62(13.06) | 22-80 | Education (years) | 13.18(2.73) | 8-19 |
| Time since diagnosis (years) | 4.06(5.28) | 0.33-31 | Largest mass diameter (cm) | 3.90(2.29) | 0.6-10.2 |
| Time since transformation (years) | 2.47(3.92) | 0.25-11 | SUVmax on PET | 16.88(10.45) | 2.1-41.1 |
|  | *N* | *%* |  | *N* | *%* |
| Diagnosis |  |  | Stage at diagnosis |  |  |
| DLBCL | 25 | 41.67 | IV | 25 | 51.02 |
| HGBL | 8 | 13.33 | III | 10 | 20.41 |
| B-ALL | 5 | 8.33 | II | 8 | 16.33 |
| DLBCL (tf FL) | 4 | 6.67 | I | 6 | 12.24 |
| DLBCL (tf MZL) | 3 | 5.00 | History of CNS disease | 8 | 13.33 |
| MCL | 3 | 5.00 | Current CNS involvement | 2 | 3.33 |
| MM | 3 | 5.00 | Current BM involvement | 13 | 21.67 |
| DLBCL (tf CLL) | 2 | 3.33 | MRD positive | 8 | 13.33 |
| HGBL (tf FL) | 2 | 3.33 | Elevated LDH (> 250) | 19 | 31.67 |
| PMBCL | 2 | 3.33 | Ferritin grade 1-2 (>ULN-5.0xULN) | 25 | 44.64 |
| FL | 1 | 1.67 | Ferritin grade 3-4 (>5.0xULN) | 13 | 23.21 |
| T-ALL | 1 | 1.67 | ECOG 0 | 41 | 68.33 |
| mRCC | 1 | 1.67 | ECOG 1 | 19 | 31.67 |
| Neurological history |  |  | Psychiatric history |  |  |
| Peripheral neuropathy | 9 | 15.00 | Symptoms of depression | 13 | 21.67 |
| Migraines | 8 | 13.33 | Symptoms of anxiety | 11 | 18.33 |
| Concussions | 3 | 5.00 | History of trauma | 3 | 5.00 |
| Benign meningioma | 1 | 1.67 | History of eating disorder | 1 | 1.67 |
| Essential tremor | 1 | 1.67 | History of psychosis | 1 | 1.67 |
| Epilepsy (seizure-free) | 1 | 1.67 | History of ADHD | 1 | 1.67 |
| Tic disorder (resolved) | 1 | 1.67 | Specific learning disorder | 1 | 1.67 |

*Note.* DLBCL = diffuse large B-cell lymphoma; HGBL = high-grade B-cell lymphoma; B-ALL = B-cell acute lymphoblastic leukaemia; tf = transformed; FL = follicular lymphoma; MZL = marginal zone lymphoma; MCL = mantle cell lymphoma; MM = multiple myeloma; CLL = chronic lymphocytic leukaemia; PMBCL = primary mediastinal large B-cell lymphoma; T-ALL = T-cell acute lymphoblastic leukaemia; mRCC = metastatic renal cell carcinoma; CNS = central nervous system; BM = bone marrow; MRD = minimal residual disease; LDH = lactate dehydrogenase; ECOG = Eastern Cooperative Oncology Group Performance Status Scale; SUVmax = maximum standardized uptake value; ADHD = attention-deficit/hyperactivity disorder; ULN = upper limit normal. Elevated ferritin was graded according to the National Cancer Institute Common Terminology Criteria for Adverse Events.

### Clinical impression

The clinical impression of the treating clinical neuropsychologist was extracted from patient files and classified as: normal cognition, subjective cognitive dysfunction, mild cognitive impairment, or moderate cognitive impairment. Table 2 in the supplementary materials describes classification categories.

**Table 2.**
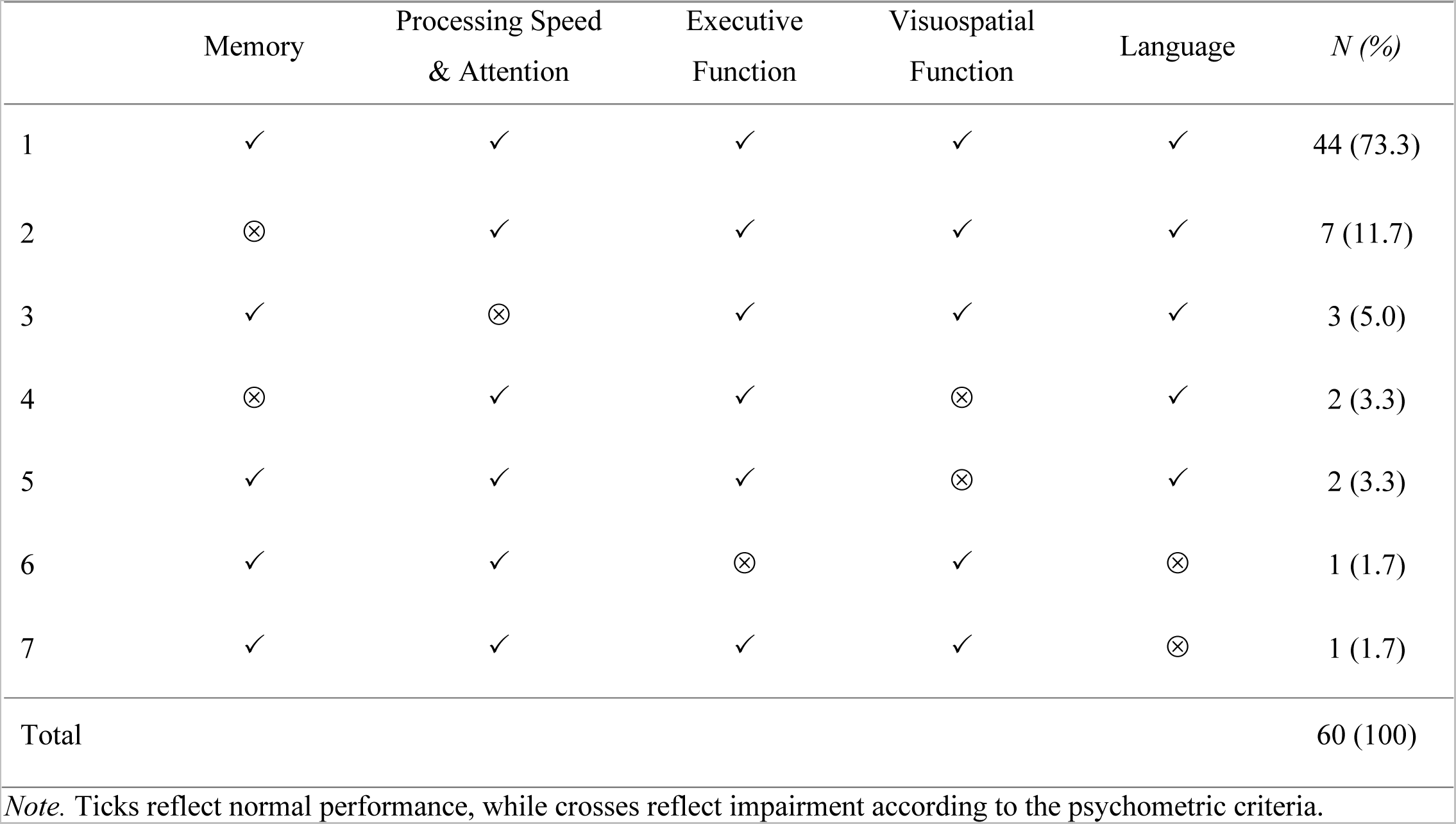
Unique patterns of impairment of 32 possible combinations

### Statistical approach

All analyses were performed using R (version 2022.12.0+353) and JASP (version 0.17.1) software packages, which are available as free and open-source programs. We reported proportions of patients impaired on each cognitive domain and each psychometric instrument, as well as the number of tests and domains most frequently impaired. An exact McNemar’s test^20^ compared impairment classification between clinical impression of the treating clinical neuropsychologist and purely psychometric criteria. Proportions of patients elevated on each SPECTRA domain were reported. The unique patterns of cognitive impairment and psychopathology were described.

The receiver operating characteristic (ROC) curve analysis examined if the MoCA or the Cognitive Concerns SPECTRA domain could identify individuals impaired according to psychometric or clinical classification. Reported statistics included areas under the curve (AUC), 95% credible intervals (CIs), sensitivity, specificity, the positive predictive value (PPV), the negative predictive value (NPV), and the best recommended threshold.

Clinicodemographic characteristics were compared between psychometrically normal and impaired patients. Bayesian independent sample t-test was used for continuous variables that satisfied assumptions of normality^21^ and homogeneity of variance^22^ and the Bayesian Mann-Whitney U test was used for those that did not.^23–25^ Effect sizes were reported by Hedges’ g^26^ and by the rank biserial correlation.^27^ Bayesian contingency table analyses were used for categorical variables.^28^ Effect sizes were reported by log odds ratios for 2x2 tables. The Bayes factors (BF_10_) were interpreted as follows: 0.33-3 as anecdotal evidence, 0.10-0.33 and 3-10 as substantial evidence for the null or for the alternative hypothesis, respectively, 0.03-0.10 and 10-30 as strong evidence in support of the null or the alternative hypothesis, respectively.^23, 29^ Missing data were dealt with via pair-wise deletion.

## Results

Sixty patients met the inclusion criteria (see Figure 1 in the supplemental materials for the exclusion process), most of whom were male (58.3%, *n*=35). Most patients underwent two prior lines of therapy (51.7%, *n*=31, range=1-8). Thirty-nine (65.0%) individuals had been exposed to CNS-penetrating chemotherapy and six (10.0%) had CNS-penetrating radiotherapy. Patients were generally examined prior to commencing conditioning lymphodepleting chemotherapy (i.e., fludarabine and cyclophosphamide; 85.0%, *n*=51). Others were examined during the chemotherapy. Most individuals were scheduled to receive commercial CAR-T products (80.0%, *n*=48), while others were planned for clinical trials. Medical and psychiatric history was obtained from medical records and reported by patients. Tables 1 outlines baseline clinical characteristics, and Table 3 in the supplementary materials specifies treatment details.

**Figure 1.**
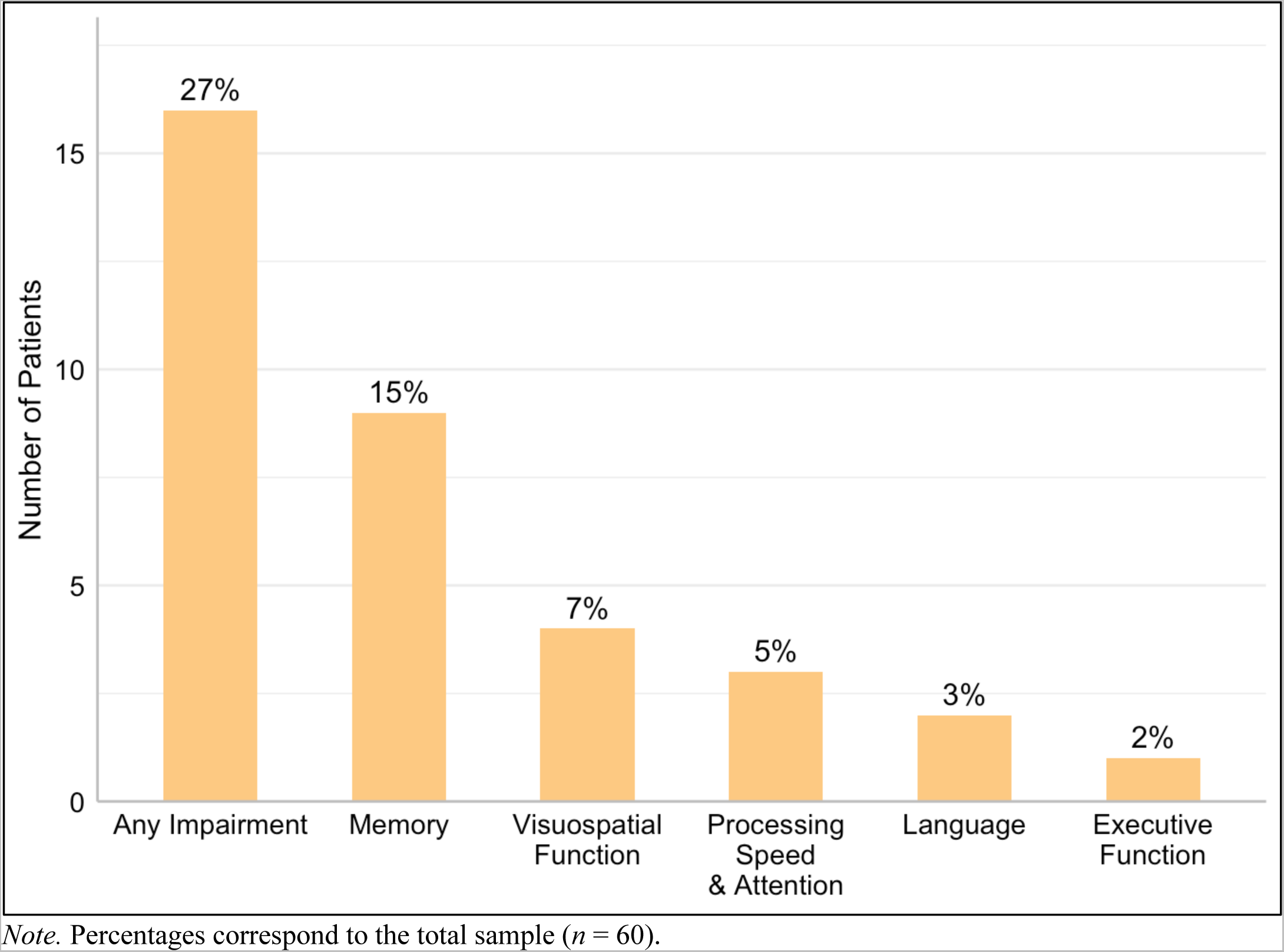
Proportion of patients impaired across cognitive domains and on any domain

**Table 3.** Unique patterns of domain elevations of 128 possible combinations

|  | Depression | Anxiety | Social<br>Anxiety | Alcohol<br>Problems | Antisocial | Psychosis | Manic<br>Activation | *N (%)* |
| --- | --- | --- | --- | --- | --- | --- | --- | --- |
| 1 | ✓ | ✓ | ✓ | ✓ | ✓ | ✓ | ✓ | 48 (84.2) |
| 2 | ✓ | ✓ | ✓ | ↑↑ | ✓ | ✓ | ✓ | 2 (3.5) |
| 3 | ✓ | ✓ | ↑↑ | ✓ | ✓ | ✓ | ✓ | 2 (3.5) |
| 4 | ✓ | ✓ | ✓ | ✓ | ✓ | ↑↑ | ✓ | 1 (1.8) |
| 5 | ✓ | ✓ | ✓ | ✓ | ↑↑ | ✓ | ✓ | 1 (1.8) |
| 6 | ✓ | ✓ | ✓ | ↑↑ | ✓ | ✓ | ↑↑ | 1 (1.8) |
| 7 | ✓ | ↑↑ | ✓ | ✓ | ✓ | ✓ | ✓ | 1 (1.8) |
| 8 | ↑↑ | ↑↑ | ✓ | ✓ | ✓ | ✓ | ✓ | 1 (1.8) |
| Total |  |  |  |  |  |  |  | 57 (100) |

*Note.* Ticks reflect normal performance, while arrow symbols reflect elevated domain scores ( $\geq 70$ ).

According to the psychometric criteria, 16 patients (26.7%) were impaired on at least one cognitive domain, with three patients (5.0%) impaired on two domains. Figure 1 depicts proportion of dysfunction across cognitive domains, and Figure 2 in the supplemental materials shows distribution of individual z-scores in each domain. Our analysis identified six unique patterns of cognitive dysfunction (see Table 2). At an instrument level, over half of our cohort (53.3%, *n*=32) was impaired on at least one psychometric test, with abnormal scores most frequent on the Trail Making Test Part B (26.7%, *n*=16). Figure 2 shows proportions of patients impaired on each instrument and on different number of tests.

**Figure 2.**
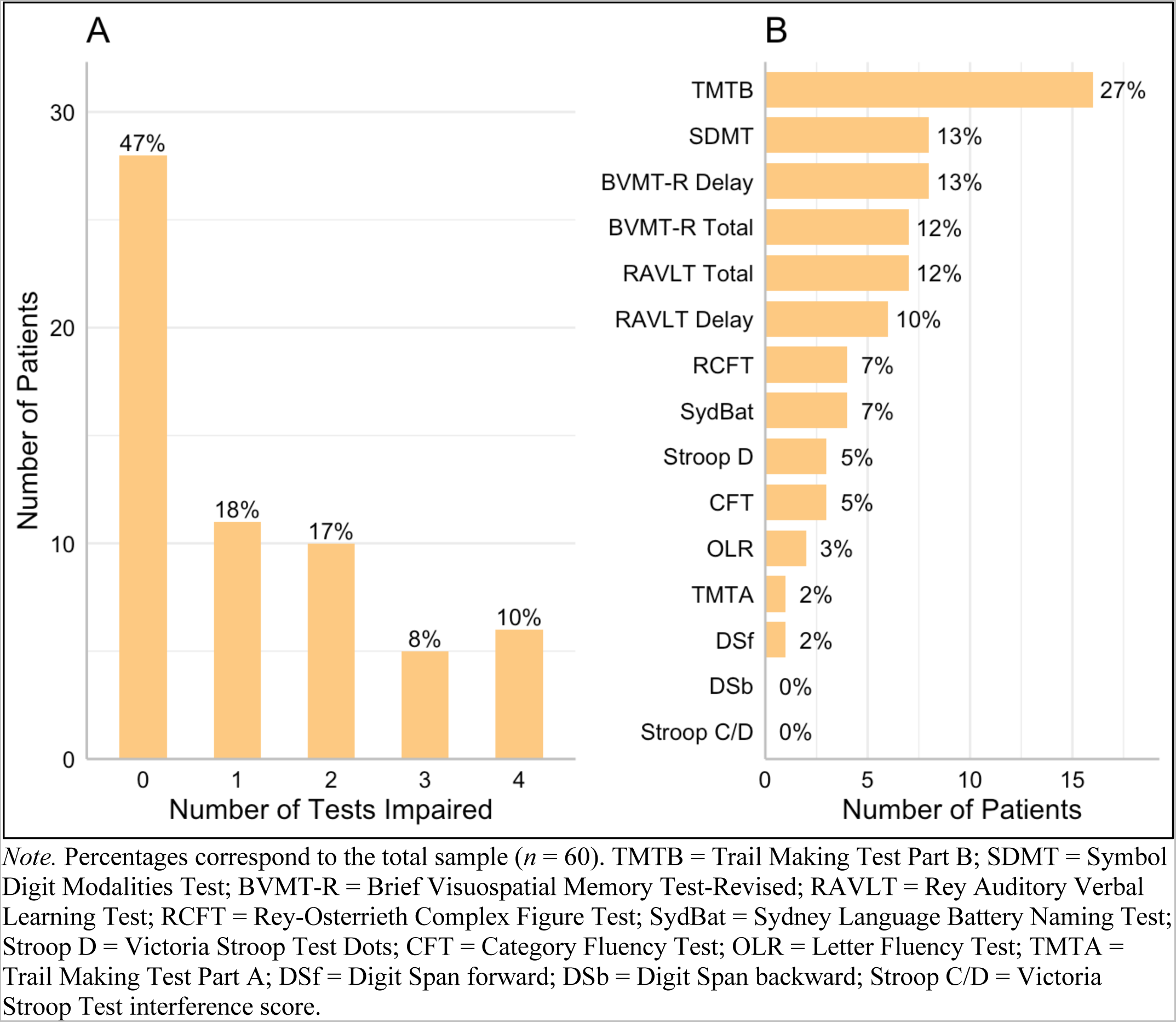
Proportion of patients impaired on different number of test and across individual tests

According to the impression of the treating clinical neuropsychologist, fifteen patients (25.0%) presented with evidence of mild cognitive impairment. Some experienced only subjective dysfunction in the absence of impairment during the examination (30.0%, *n*=18). No patient met the criteria for moderate impairment. To compare clinical impression with psychometric performance, patients with normal cognition and those with only subjective dysfunction were classified as ‘normal’, while the remainder of the cohort was classified as ‘impaired’. Although prevalence of cognitive impairment was similar for both approaches, four psychometrically normal individuals (6.7%) were impaired according to the treating clinician. In contrast, five patients (8.3%) who were normal based on clinical impression, were psychometrically impaired. An exact McNemar’s test confirmed significant discordance in classification of individual cases, *χ*^2^(1, N=60)=15.37, *p*<.001.

Forty-six patients (76.7%) completed the MoCA, of whom 10 received abnormal scores <26.^11^ Of this subset, nine patients (19.6%) were impaired based on the psychometric criteria and six (13.0%) based on the clinical impression. The ROC curves in Figure 3.A compare total MoCA scores against psychometric performance (*AUC*=0.61, 95% CIs [0.42– 0.81]) and against clinical classification (*AUC*=0.56, 95% CIs [0.31–0.81]). Compared to the psychometric criteria, the MoCA demonstrated sensitivity of 0.33, specificity of 0.81, the PPV of 0.30, and the NPV of 0.83, which approximated prevalence of normal cognition in our cohort. Compared to the clinical classification, the MoCA had sensitivity of 0.33, specificity of 0.80, the PPV of 0.20, and the NPV of 0.89. The recommended best threshold for an abnormal MoCA score was <28 for both psychometric (sensitivity=0.78, specificity=0.51, PPV=0.28, NPV=0.91) and clinical (sensitivity=0.67, specificity=0.48, PPV=0.16, NPV=0.91) comparison. In our cohort, 25 patients (54.3%) received a MoCA score <28.

**Figure 3.**
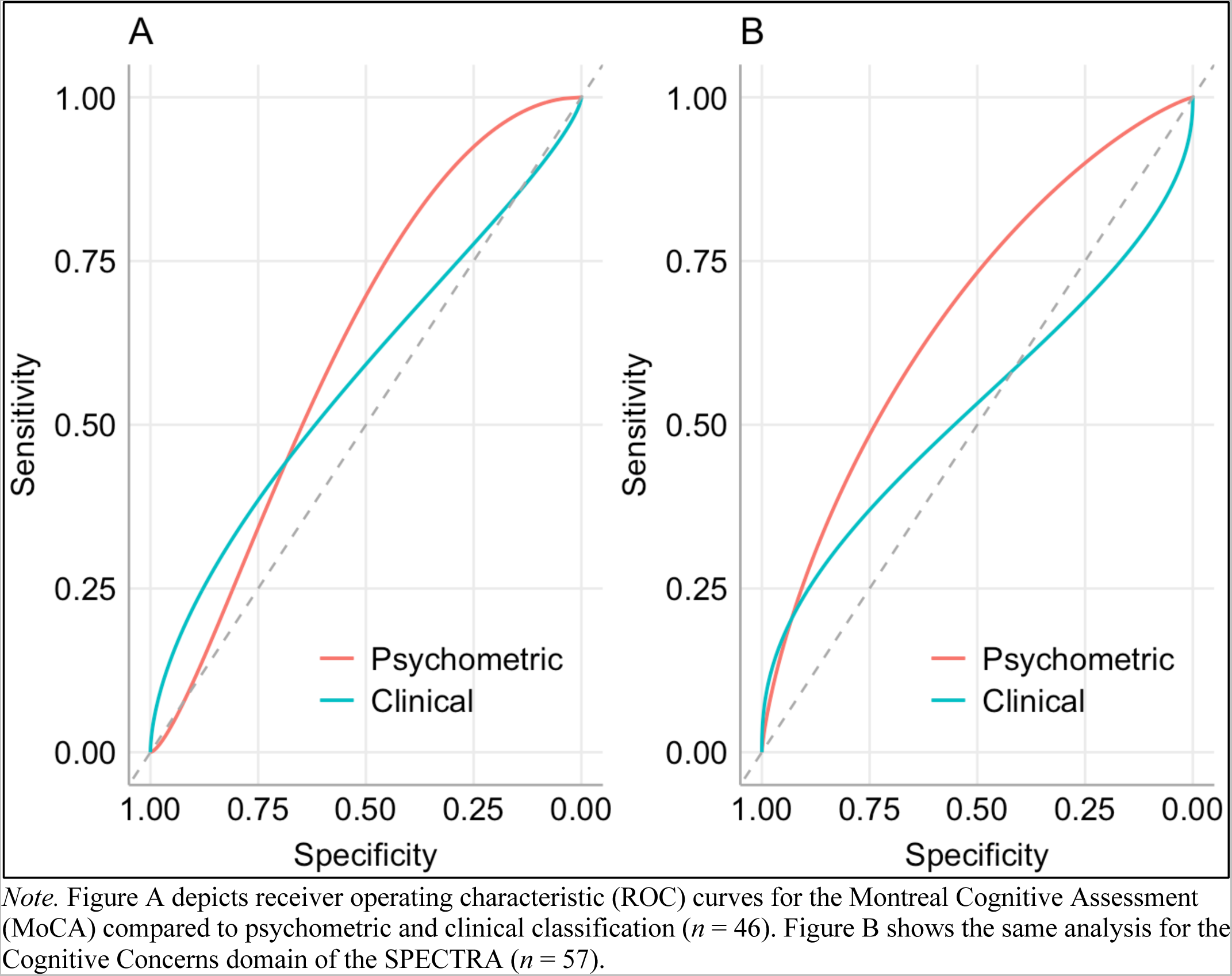
ROC curves of the MoCA and the Cognitive Concerns performance against psychometric and clinical classification of cognitive impairment

Fifty-seven participants (95.0%) completed the SPECTRA questionnaire. Nine patients (15.8%) were elevated (t-score≥70)^17^ on at least one symptom domain, with two patients (3.5%) elevated on two domains. Figure 4 shows proportions of patients elevated on each SPECTRA domain. There were seven unique patterns of psychopathology (see Table 3).

**Figure 4.**
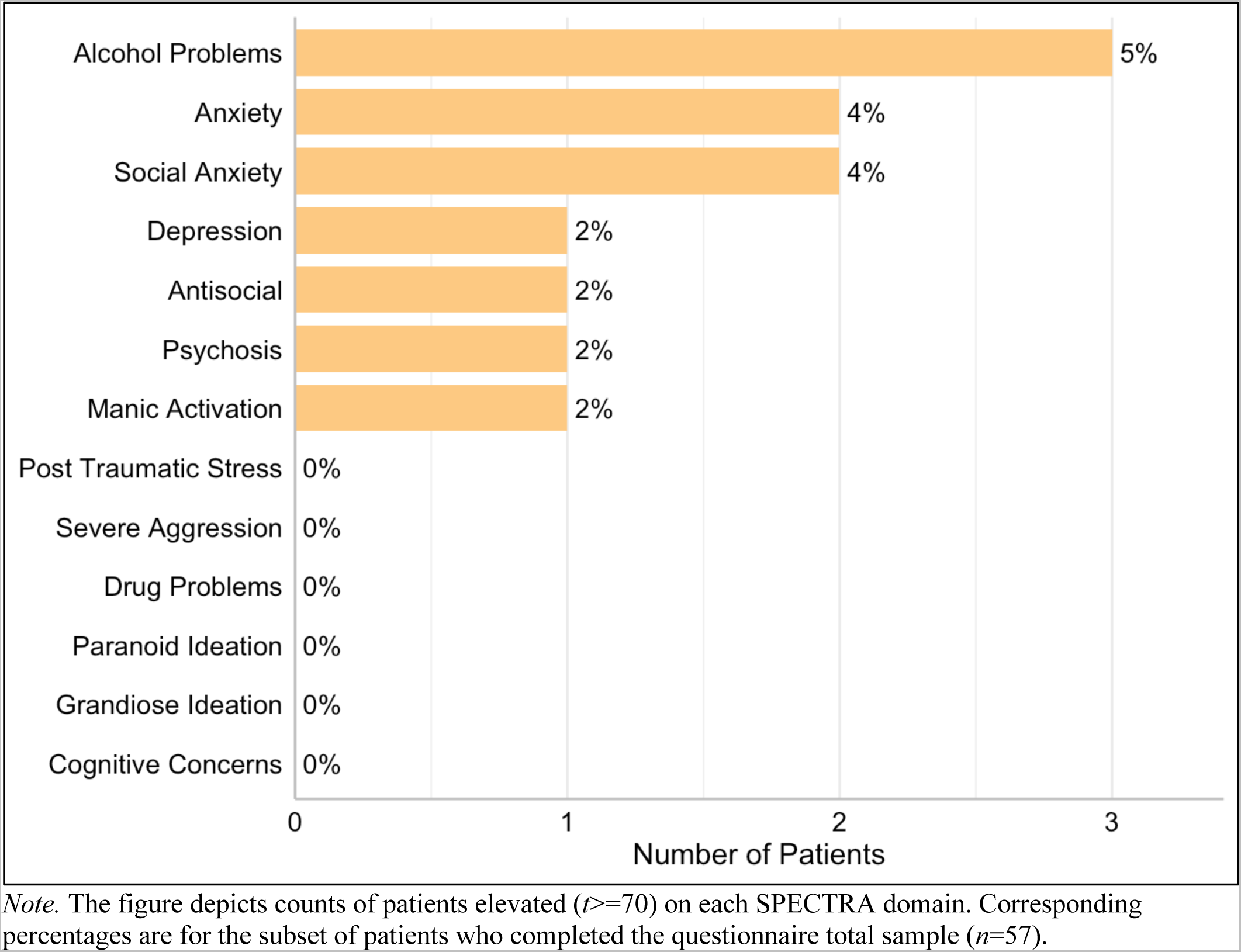
Proportion of patients elevated on each SPECTRA domain

The ROC curve analysis examined whether responses on the Cognitive Concerns SPECTRA domain adequately distinguished between patients who were impaired on psychometric or clinical grounds (see Figure 3.B). Of this subset, 14 patients (24.6%) were impaired based on the psychometric criteria and 14 (24.6%) were impaired based on clinical impression. No individuals, however, were elevated on the Cognitive Concerns. Comparing the SPECTRA against the psychometric criteria, the AUC was 0.64 (95% CIs [0.47–0.81]). The AUC was 0.54 (95% CIs [0.29–0.78]) for the comparison against clinical classification. Since no patients obtained a t-score ≥70 on the Cognitive Concerns, statistics are not reported for this threshold. The recommended best threshold was ≥59.5 for both psychometric (sensitivity=0.36, specificity=0.93, PPV=0.63, NPV=0.82) and clinical (sensitivity=0.22, specificity=0.96, PPV=0.67, NPV=0.78) comparison. In our cohort, eight patients (14.0%) received a t-score ≥59.5 on the Cognitive Concerns, however, placing a threshold for psychopathology 0.95z above the population mean is not statistically or clinically feasible.

Patients who were psychometrically impaired were compared on clinicodemographic characteristics to those with normal performance (see Table 4). Bayesian Mann-Whitney U test demonstrated substantial evidence that impaired patients were younger (*M*=50.00, *SD*=14.50) than those cognitively intact (*M*=60.39, *SD*=11.45), *U*=513.50, *p*=.007, *r_rb_*=0.46, *BF_10_*=7.76. Bayesian contingency table analyses indicated substantial evidence for group difference on diagnosis, with B-cell lymphoblastic leukaemia patients more frequently impaired, *χ*^2^(3, N=60)=9.44, *p*=.024, *v*=0.40, *BF_10_*=9.30. Substantial evidence indicated that patients with bone marrow involvement were more likely to be impaired, *χ*^2^(1, N=55)=5.06, *p*=.037, *OR*=1.42, *BF_10_*=5.18, as were those with reported history of anxiety, *χ*^2^(1, N=60)=5.35, *p*=.053, *OR*=1.51, *BF_10_*=4.85. Very strong evidence indicated that individuals elevated on any SPECTRA domain were more likely to have impaired psychometric performance, *χ*^2^(1, N=57)=10.23, *p*=.004, *OR*=2.25, *BF_10_*=31.30. The estimated Bayes factors showed some evidence that the groups were equivalent in education and in years since diagnosis. There was no significant difference between groups on any other characteristics, however, the evidence was considered anecdotal.

**Table 4.**
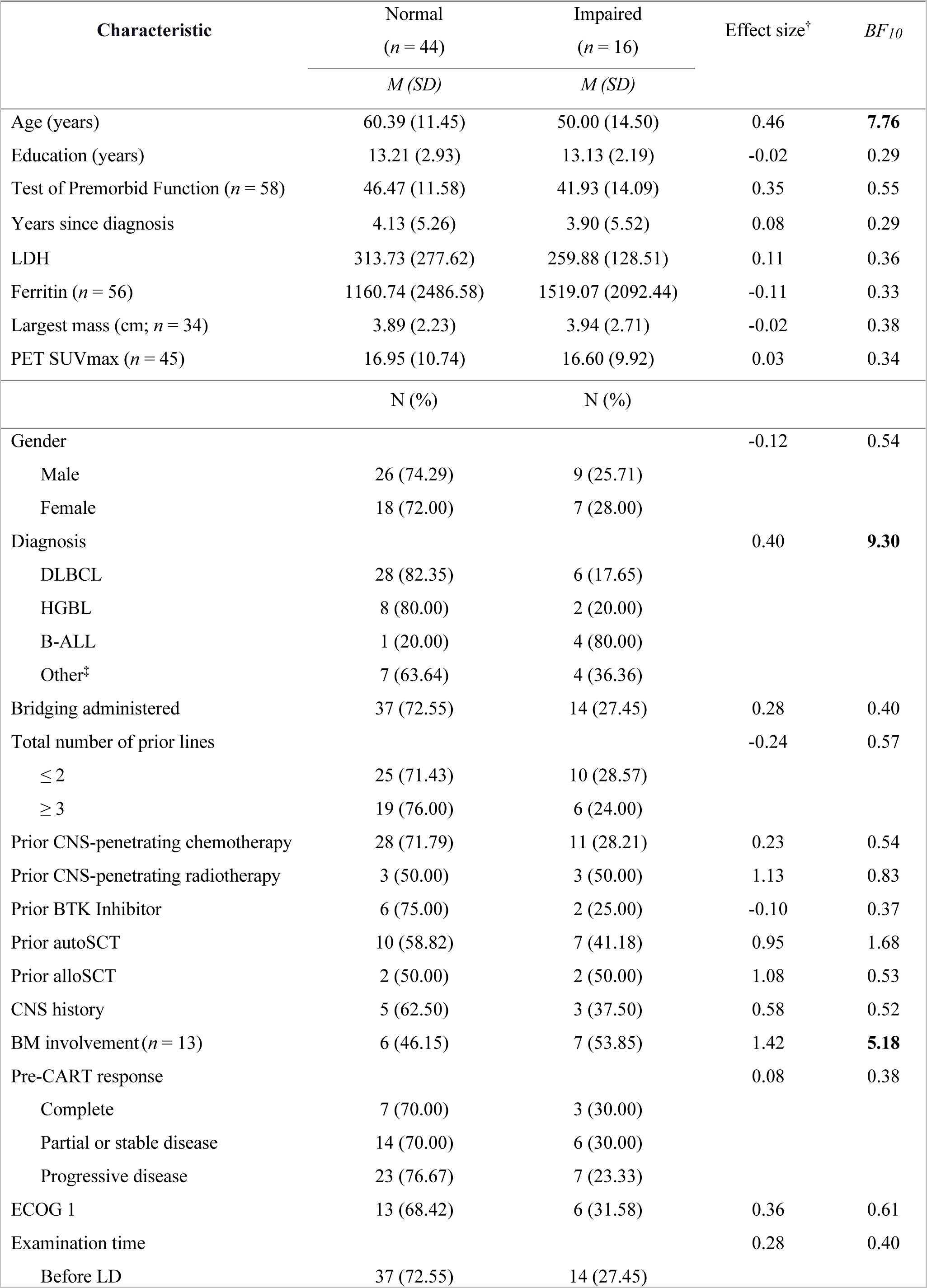
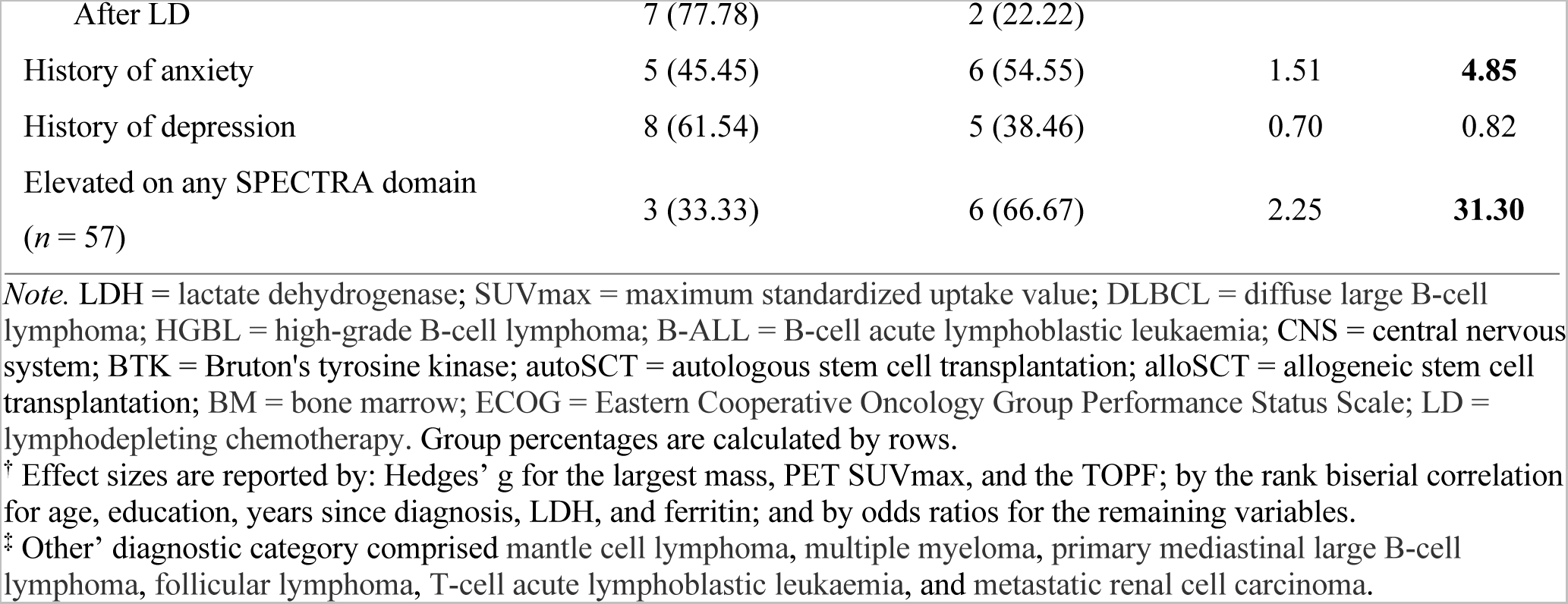
Clinicodemographic characteristics of patients grouped by psychometric impairment

| Characteristic | Normal<br>(*n* = 44) | Impaired<br>(*n* = 16) | Effect size<sup>†</sup> | *BF*<sub>10</sub> |
| --- | --- | --- | --- | --- |
|  | *M* (*SD*) | *M* (*SD*) |  |  |
| Age (years) | 60.39 (11.45) | 50.00 (14.50) | 0.46 | **7.76** |
| Education (years) | 13.21 (2.93) | 13.13 (2.19) | -0.02 | 0.29 |
| Test of Premorbid Function (*n* = 58) | 46.47 (11.58) | 41.93 (14.09) | 0.35 | 0.55 |
| Years since diagnosis | 4.13 (5.26) | 3.90 (5.52) | 0.08 | 0.29 |
| LDH | 313.73 (277.62) | 259.88 (128.51) | 0.11 | 0.36 |
| Ferritin (*n* = 56) | 1160.74 (2486.58) | 1519.07 (2092.44) | -0.11 | 0.33 |
| Largest mass (cm; *n* = 34) | 3.89 (2.23) | 3.94 (2.71) | -0.02 | 0.38 |
| PET SUVmax (*n* = 45) | 16.95 (10.74) | 16.60 (9.92) | 0.03 | 0.34 |
|  | N (%) | N (%) |  |  |
| Gender |  |  | -0.12 | 0.54 |
| Male | 26 (74.29) | 9 (25.71) |  |  |
| Female | 18 (72.00) | 7 (28.00) |  |  |
| Diagnosis |  |  | 0.40 | **9.30** |
| DLBCL | 28 (82.35) | 6 (17.65) |  |  |
| HGBL | 8 (80.00) | 2 (20.00) |  |  |
| B-ALL | 1 (20.00) | 4 (80.00) |  |  |
| Other<sup>‡</sup> | 7 (63.64) | 4 (36.36) |  |  |
| Bridging administered | 37 (72.55) | 14 (27.45) | 0.28 | 0.40 |
| Total number of prior lines |  |  | -0.24 | 0.57 |
| ≤ 2 | 25 (71.43) | 10 (28.57) |  |  |
| ≥ 3 | 19 (76.00) | 6 (24.00) |  |  |
| Prior CNS-penetrating chemotherapy | 28 (71.79) | 11 (28.21) | 0.23 | 0.54 |
| Prior CNS-penetrating radiotherapy | 3 (50.00) | 3 (50.00) | 1.13 | 0.83 |
| Prior BTK Inhibitor | 6 (75.00) | 2 (25.00) | -0.10 | 0.37 |
| Prior autoSCT | 10 (58.82) | 7 (41.18) | 0.95 | 1.68 |
| Prior alloSCT | 2 (50.00) | 2 (50.00) | 1.08 | 0.53 |
| CNS history | 5 (62.50) | 3 (37.50) | 0.58 | 0.52 |
| BM involvement (*n* = 13) | 6 (46.15) | 7 (53.85) | 1.42 | **5.18** |
| Pre-CART response |  |  | 0.08 | 0.38 |
| Complete | 7 (70.00) | 3 (30.00) |  |  |
| Partial or stable disease | 14 (70.00) | 6 (30.00) |  |  |
| Progressive disease | 23 (76.67) | 7 (23.33) |  |  |
| ECOG 1 | 13 (68.42) | 6 (31.58) | 0.36 | 0.61 |
| Examination time |  |  | 0.28 | 0.40 |
| Before LD | 37 (72.55) | 14 (27.45) |  |  |
| After LD | 7 (77.78) | 2 (22.22) |  |  |
| History of anxiety | 5 (45.45) | 6 (54.55) | 1.51 | **4.85** |
| History of depression | 8 (61.54) | 5 (38.46) | 0.70 | 0.82 |
| Elevated on any SPECTRA domain<br>(*n* = 57) | 3 (33.33) | 6 (66.67) | 2.25 | **31.30** |

*Note.* LDH = lactate dehydrogenase; SUVmax = maximum standardized uptake value; DLBCL = diffuse large B-cell lymphoma; HGBL = high-grade B-cell lymphoma; B-ALL = B-cell acute lymphoblastic leukaemia; CNS = central nervous system; BTK = Bruton's tyrosine kinase; autoSCT = autologous stem cell transplantation; alloSCT = allogeneic stem cell transplantation; BM = bone marrow; ECOG = Eastern Cooperative Oncology Group Performance Status Scale; LD = lymphodepleting chemotherapy. Group percentages are calculated by rows.
<sup>†</sup> Effect sizes are reported by: Hedges' *g* for the largest mass, PET SUVmax, and the TOPF; by the rank biserial correlation for age, education, years since diagnosis, LDH, and ferritin; and by odds ratios for the remaining variables.
<sup>‡</sup> Other' diagnostic category comprised mantle cell lymphoma, multiple myeloma, primary mediastinal large B-cell lymphoma, follicular lymphoma, T-cell acute lymphoblastic leukaemia, and metastatic renal cell carcinoma.

## Discussion

As anticipated, a substantial proportion of patients presented with cognitive impairment based on either the impression of the treating clinical neuropsychologist or solely on psychometric grounds. Prevalence of impairment notably exceeded dysfunction expected in the normal population (6.7% for threshold of −1.5z). Impaired performance was observed across all cognitive domains (i.e., memory, visuospatial function, processing speed and attention, language, and executive function), with most impairments observed on the memory domain. Abnormal psychometric performance was more likely in individuals who were younger, had B-cell acute lymphoblastic leukaemia (B-ALL), had bone marrow involvement, reported history of anxiety, or had evidence of psychopathology. The increased risk in the B- ALL cohort might not be surprising given the intensity of CNS directed therapy used in the standard-of-care regimens to treat this condition. There was no single psychometric profile, with six unique patterns of cognitive dysfunction identified. Such variability supports the diverse psychometric battery approach to adequately capture impairment in this cohort. The findings are comparable with descriptions of cancer-related cognitive impairment in the literature. For example, a review by Janelsins et al.^30^ found that approximately 30% of oncology patients experienced cognitive dysfunction in a non-acute treatment setting. Impairment was common in memory, executive function, speed and attention. Clinical manifestations of ICANS are commonly described to involve dysfunction in attention, memory, and language.^31, 32^ Thus, patients with unexamined cognition prior to CAR-T are at risk of misdiagnosis post-infusion.

There was significant discordance in impairment classification between clinical impression and the psychometric criteria. Researchers often adopt a psychometric approach for good reliability and lower resource requirement. There might, however, be aspects of clinical activity that are not replicated by a solely psychometric model. For example, secondary factors, such as pain, fatigue, poor sleep, and stress, are prevalent in the cancer population^33–35^ and can temporarily disrupt cognitive function leading to impaired psychometric performance.^34, 36^ A probabilistic distinction between primary and secondary dysfunction is commonly made in clinical practice, which is difficult to capture psychometrically. Although our findings do not determine superiority of the clinical approach, further inquiry is warranted to empirically investigate the difference between primary and secondary impairment, and its relevance for clinical management.

Resources in cancer centres can be scarce, limiting the number of patients who can complete specialist cognitive examination pre-CAR-T. We analysed whether screening with a measure of objective cognitive impairment (the MoCA) or a measure of psychometric self-report of cognitive function (the Cognitive Concerns SPECTRA domain) could adequately detect individuals with suspected cognitive impairment to be prioritised for further examination. Unfortunately, both instruments performed poorly against psychometric performance and clinical classification.

The demonstrated futility of cognitive screening contrasts with De Roeck et al.,^12^ who found the MoCA useful in detecting cognitive impairment in early presentation of dementia of the Alzheimer’s type. The instrument, however, lacks items targeting cognitive functions most frequently impaired in our cohort (e.g., processing speed, memory, and higher order attention). Thus, the MoCA might not detect relatively subtle dysfunction specific to these patients. The absence of significant concerns on the self-report measure of cognitive function was also somewhat unexpected. Subjective cognitive dysfunction has been frequently reported in haematology patients.^15, 37^ The strong emphasis on daily function in the Cognitive Concerns domain of the SPECTRA^17^ could explain this discordance. Eligibility for CAR-T therapy requires adequate functional status, naturally excluding individuals with functional decline from our cohort. Cognitive complaint is phenomenologically broader than aspects captured by common self-report questionnaires.^16^ Future work could consider investigating semiology of a cognitive complaint in CAR-T patients to capture the complexity of their subjective experience. Further research into development and validation of appropriate screening instruments for CAR-T patients is also a meaningful endeavour.

A substantial proportion of patients were elevated on the self-report measures of psychopathology, with seven unique patters identified. Symptoms spanned beyond depression and anxiety, commonly described in cancer patients,^9, 10^ and included alcohol problems, psychosis, antisocial behaviours, and manic activation domains. Interestingly, the prevalence of clinically elevated depression and anxiety was lower than anticipated in oncology population.^9, 10^ Presumably, our cohort could be more robust than recently diagnosed and treatment naïve patients. The average time since diagnosis was four years, and all patients received prior systemic therapy. The time taken to adjust to the diagnosis, past treatment experience, and relatively good functional status might serve as protective factors. The specific structure of CAR-T program, coupled with education and support provided at our centre, might also reduce anxiety by shifting focus to more immediate practical matters.^38^ Consistent with the literature, patients with evidence of psychopathology were 2.25 times more likely to be cognitively impaired.^39^ Our findings emphasise the complexity of pre-CAR-T emotional status and highlight the value in consideration of associated cognitive contribution.

The current study was the first to prospectively and systematically investigate baseline cognitive and psychological status in CAR-T patients. We evaluated cognition with a comprehensive battery of validated and reliable psychometric instruments, as well as the impression of an experienced clinical neuropsychologist. Targeted cognitive domains aligned with recommendations from the ICCTF^18^ and included those commonly impaired in cancer patients. Our methodology was derived from routine clinical practice at our haematology service, facilitating practically meaningful interpretation of the findings. There are, however, several limitations to consider. The visuospatial function domain comprised a single psychometric measure. Incorporating additional instruments could improve reliability.

Interpretation of responses on the self-report measures of psychopathology utilized a threshold for clinically significant presentation.^17^ Such approach captures only those with severe psychopathology. A more nuanced evaluation of psychological status in CAR-T patients could better capture a broad spectrum of possible symptoms. Finally, comparing clinicodemographic features between psychometrically normal and impaired individuals was largely an exploratory exercise, with limited power for many characteristics. Further study is needed to identify predictors of cognitive dysfunction in this cohort.

## Conclusion

The current study provides a unique insight into cognition of patients planned for CAR-T therapy. It forms a comprehensive baseline for future investigation of neurocognitive manifestation of ICANS and its recovery trajectory. We have determined that presently, a cognitive screening approach is not sufficient, and specialist cognitive evaluation is necessary to accurately detect impairment in this population. Further study is encouraged to identify a valid screening instrument and to adequately capture a subjective cognitive complaint in this cohort. Understanding the nature of cognitive dysfunction and psychopathology in these patients will advance patient care, including counselling and referral pathways.

## Data Availability

Anonymised data, and the standardized proforma used to extract information on patient demographics and clinical characteristics, can be shared on reasonable request from qualified investigators.

## Acknowledgements

We acknowledge and thank CAR-T patients and their families for their participation. We also thank medical and administrative teams at Peter MacCallum Cancer Centre for their patient care and logistical contribution to this study. All authors substantially contributed to this work, with specific contributions outline in the Authorship Statement below.

## Authorship statement

Valeriya Kuznetsova, Charles B Malpas, Hannah Rosenfeld, Mary Ann Anderson, Michael Dickinson, Tomas Kalincik, Simon J Harrison, and Izanne Roos contributed to research design. Data was collected by Valeriya Kuznetsova, Hannah Rosenfeld, Carmela Sales, Samantha van der Linde, Mark Dowling, and Stefanie Roberts. Valeriya Kuznetsova, Charles B Malpas, Tomas Kalincik, Fiore D’Aprano, and Samantha M Loi were involved in analyses and/or interpretation of the findings. All authors contributed to the final manuscript and approved it for submission.

## References

1. Rice J, Nagle S, Randall J, Hinson HE. Chimeric Antigen Receptor T Cell-Related Neurotoxicity: Mechanisms, Clinical Presentation, and Approach to Treatment. Curr Treat Options Neurol. 2019;21(8):40. doi:10.1007/s11940-019-0580-3

2. Siegler EL, Kenderian SS. Neurotoxicity and Cytokine Release Syndrome After Chimeric Antigen Receptor T Cell Therapy: Insights Into Mechanisms and Novel Therapies. Front Immunol. 2020;11. Accessed May 29, 2023. https://www.frontiersin.org/articles/10.3389/fimmu.2020.01973

3. Herr MM, Chen GL, Ross M, et al. Identification of Neurotoxicity after Chimeric Antigen Receptor (CAR) T Cell Infusion without Deterioration in the Immune Effector Cell-Associated Encephalopathy (ICE) Score. Biol Blood Marrow Transplant. 2020;26(11):e271–e274. doi:10.1016/j.bbmt.2020.07.031

4. Mayo SJ, Lustberg M, M. Dhillon H, et al. Cancer-related cognitive impairment in patients with non-central nervous system malignancies: an overview for oncology providers from the MASCC Neurological Complications Study Group. Support Care Cancer. 2021;29(6):2821–2840. doi:10.1007/s00520-020-05860-9

5. Joly F, Castel H, Tron L, Lange M, Vardy J. Potential Effect of Immunotherapy Agents on Cognitive Function in Cancer Patients. JNCI J Natl Cancer Inst. 2020;112(2):123–127. doi:10.1093/jnci/djz168

6. Koll TT, Sheese AN, Semin J, et al. Screening for cognitive impairment in older adults with hematological malignancies using the Montreal Cognitive Assessment and neuropsychological testing. J Geriatr Oncol. 2020;11(2):297–303. doi:10.1016/j.jgo.2019.11.007

7. Schroyen G, Meylaers M, Deprez S, et al. Prevalence of leukoencephalopathy and its potential cognitive sequelae in cancer patients. J Chemother. 2020;32(7):327–343. doi:10.1080/1120009X.2020.1805239

8. Murrough JW, Iacoviello B, Neumeister A, Charney DS, Iosifescu DV. Cognitive dysfunction in depression: Neurocircuitry and new therapeutic strategies. Neurobiol Learn Mem. 2011;96(4):553–563. doi:10.1016/j.nlm.2011.06.006

9. Murri MB, Caruso R, Christensen AP, Folesani F, Nanni MG, Grassi L. The facets of psychopathology in patients with cancer: Cross-sectional and longitudinal network analyses. J Psychosom Res. 2023;165:111139. doi:10.1016/j.jpsychores.2022.111139

10. Clinton-McHarg T, Carey M, Sanson-Fisher R, Tzelepis F, Bryant J, Williamson A. Anxiety and depression among haematological cancer patients attending treatment centres: Prevalence and predictors. J Affect Disord. 2014;165:176–181. doi:10.1016/j.jad.2014.04.072

11. Nasreddine ZS, Phillips NA, Bédirian V, et al. The Montreal Cognitive Assessment, MoCA: A Brief Screening Tool For Mild Cognitive Impairment. J Am Geriatr Soc. 2005;53(4):695–699. doi:10.1111/j.1532-5415.2005.53221.x

12. De Roeck EE, De Deyn PP, Dierckx E, Engelborghs S. Brief cognitive screening instruments for early detection of Alzheimer’s disease: a systematic review. Alzheimers Res Ther. 2019;11(1):21. doi:10.1186/s13195-019-0474-3

13. Kotb MG, Soliman AER, Ibrahim RI, Said RMM, El Din MMW. Chemotherapy-induced cognitive impairment in hematological malignancies. Egypt J Neurol Psychiatry Neurosurg. 2019;55(1):56. doi:10.1186/s41983-019-0104-9

14. Hutchinson AD, Thompson E, Loft N, Lewis I, Wilson C, Yong ASM. Cognitive late effects following allogeneic stem cell transplantation in haematological cancer patients. Eur J Cancer Care (Engl). 2021;30(5):e13448. doi:10.1111/ecc.13448

15. Maillet D, Belin C, Moroni C, et al. Evaluation of mid-term (6-12 months) neurotoxicity in B-cell lymphoma patients treated with CAR T cells: a prospective cohort study. Neuro-Oncol. 2021;23(9):1569–1575. doi:10.1093/neuonc/noab077

16. Buckley RF, Ellis KA, Ames D, et al. Phenomenological characterization of memory complaints in preclinical and prodromal Alzheimer’s disease. Neuropsychology. 2015;29:571–581. doi:10.1037/neu0000156

17. Blais MA, Sinclair SJ. Introduction to the SPECTRA: Indices of Psychopathology:

18. Wefel JS, Vardy J, Ahles T, Schagen SB. International Cognition and Cancer Task Force recommendations to harmonise studies of cognitive function in patients with cancer. Lancet Oncol. 2011;12(7):703–708. doi:10.1016/S1470-2045(10)70294-1

19. Jak AJ, Bondi MW, Delano-Wood L, et al. Quantification of Five Neuropsychological Approaches to Defining Mild Cognitive Impairment. Am J Geriatr Psychiatry. 2009;17(5):368–375. doi:10.1097/JGP.0b013e31819431d5

20. McNemar Q. Note on the sampling error of the difference between correlated proportions or percentages. Psychometrika. 1947;12(2):153–157. doi:10.1007/BF02295996

21. Shapiro SS, Wilk MB. An Analysis of Variance Test for Normality (Complete Samples). Biometrika. 1965;52(3/4):591–611. doi:10.2307/2333709

22. Brown MB, Forsythe AB. Robust Tests for the Equality of Variances. J Am Stat Assoc. 1974;69(346):364–367. doi:10.1080/01621459.1974.10482955

23. Jeffreys H. The Theory of Probability. OUP Oxford; 1998.

24. Wagenmakers EJ. A practical solution to the pervasive problems ofp values. Psychon Bull Rev. 2007;14(5):779–804. doi:10.3758/BF03194105

25. Nachar N. The Mann-Whitney U: A Test for Assessing Whether Two Independent Samples Come from the Same Distribution. Tutor Quant Methods Psychol. 2008;4(1):13–20. doi:10.20982/tqmp.04.1.p013

26. Hedges LV. Distribution Theory for Glass’s Estimator of Effect Size and Related Estimators. J Educ Stat. 1981;6(2):107–128. doi:10.2307/1164588

27. Cureton EE. Rank-biserial correlation. Psychometrika. 1956;21(3):287–290. doi:10.1007/BF02289138

28. Gunel E, Dickey J. Bayes Factors for Independence in Contingency Tables. Biometrika. 1974;61(3):545–557. doi:10.2307/2334738

29. Jarosz A, Wiley J. What Are the Odds? A Practical Guide to Computing and Reporting Bayes Factors. J Probl Solving. 2014;7(1). doi:10.7771/1932-6246.1167

30. Janelsins MC, Kesler SR, Ahles TA, Morrow GR. Prevalence, mechanisms, and management of cancer-related cognitive impairment. Int Rev Psychiatry. 2014;26(1):102–113. doi:10.3109/09540261.2013.864260

31. Santomasso B, Bachier C, Westin J, Rezvani K, Shpall EJ. The Other Side of CAR T-Cell Therapy: Cytokine Release Syndrome, Neurologic Toxicity, and Financial Burden. Am Soc Clin Oncol Educ Book. 2019;(39):433–444. doi:10.1200/EDBK_238691

32. Tallantyre EC, Evans NA, Parry-Jones J, Morgan MPG, Jones CH, Ingram W. Neurological updates: neurological complications of CAR-T therapy. J Neurol. 2021;268(4):1544–1554. doi:10.1007/s00415-020-10237-3

33. Allart-Vorelli P, Porro B, Baguet F, Michel A, Cousson-Gélie F. Haematological cancer and quality of life: a systematic literature review. Blood Cancer J. 2015;5(4):e305–e305. doi:10.1038/bcj.2015.29

34. Drijver AJ, Oort Q, Otten R, Reijneveld JC, Klein M. Is poor sleep quality associated with poor neurocognitive outcome in cancer survivors? A systematic review. J Cancer Surviv. Published online May 2, 2022. doi:10.1007/s11764-022-01213-z

35. Priscilla D, Hamidin A, Azhar MohdZ, Noorjan KON, Salmiah MohdS, Bahariah K. Assessment of Depression and Anxiety in Haematological Cancer Patients and their Relationship with Quality of Life. East Asian Arch Psychiatry. 2011;21(3):108–114. Accessed May 30, 2023. https://search.ebscohost.com/login.aspx?direct=true&AuthType=sso&db=a9h&AN=66803220&site=ehost-live&custid=s2775460

36. van der Leeuw G, Eggermont LHP, Shi L, et al. Pain and Cognitive Function Among Older Adults Living in the Community. J Gerontol Ser A. 2016;71(3):398–405. doi:10.1093/gerona/glv166

37. Williams AM, Zent CS, Janelsins MC. What is known and unknown about chemotherapy-related cognitive impairment in patients with haematological malignancies and areas of needed research. Br J Haematol. 2016;174(6):835–846. doi:10.1111/bjh.14211

38. Fergus TA, Wheless NE. The attention training technique causally reduces self-focus following worry provocation and reduces cognitive anxiety among self-focused individuals. J Behav Ther Exp Psychiatry. 2018;61:66–71. doi:10.1016/j.jbtep.2018.06.006

39. Abramovitch A, Short T, Schweiger A. The C Factor: Cognitive dysfunction as a transdiagnostic dimension in psychopathology. Clin Psychol Rev. 2021;86:102007. doi:10.1016/j.cpr.2021.102007

