## Supplementary material for "Complexity of baseline cognitive and psychological status in haematology patients planned for chimeric antigen receptor T-cell therapy"

**Figure 1**

*CONSORT flowchart of participants*

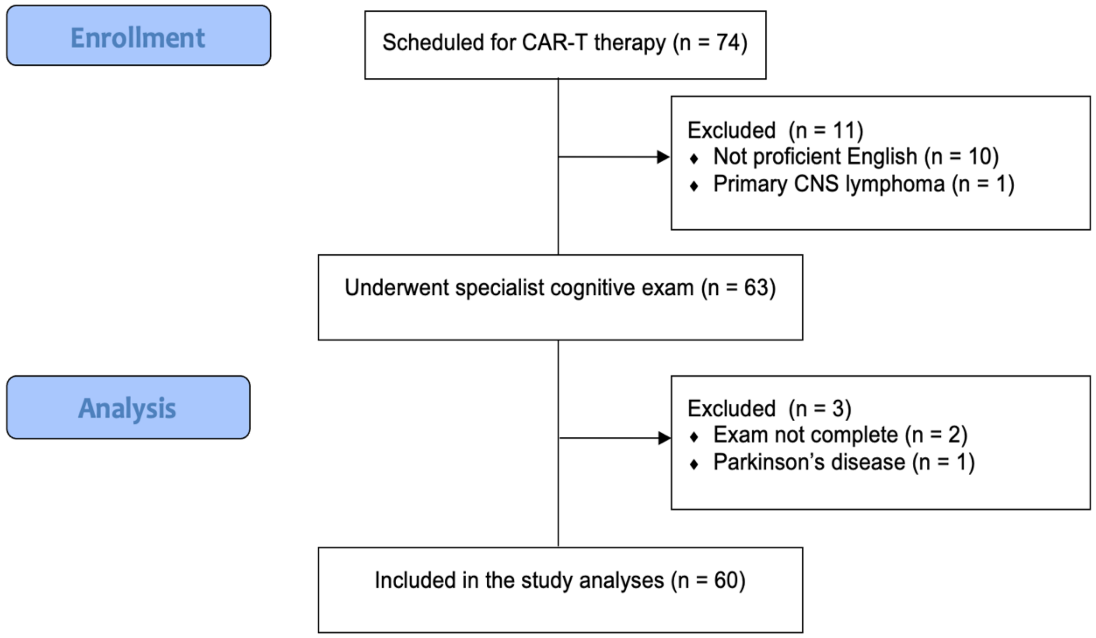


*Note.* CAR-T = chimeric antigen receptor T-cell therapy; CNS = central nervous system.

**Figure 2**

*Individual patients' z-scores across each cognitive domain*

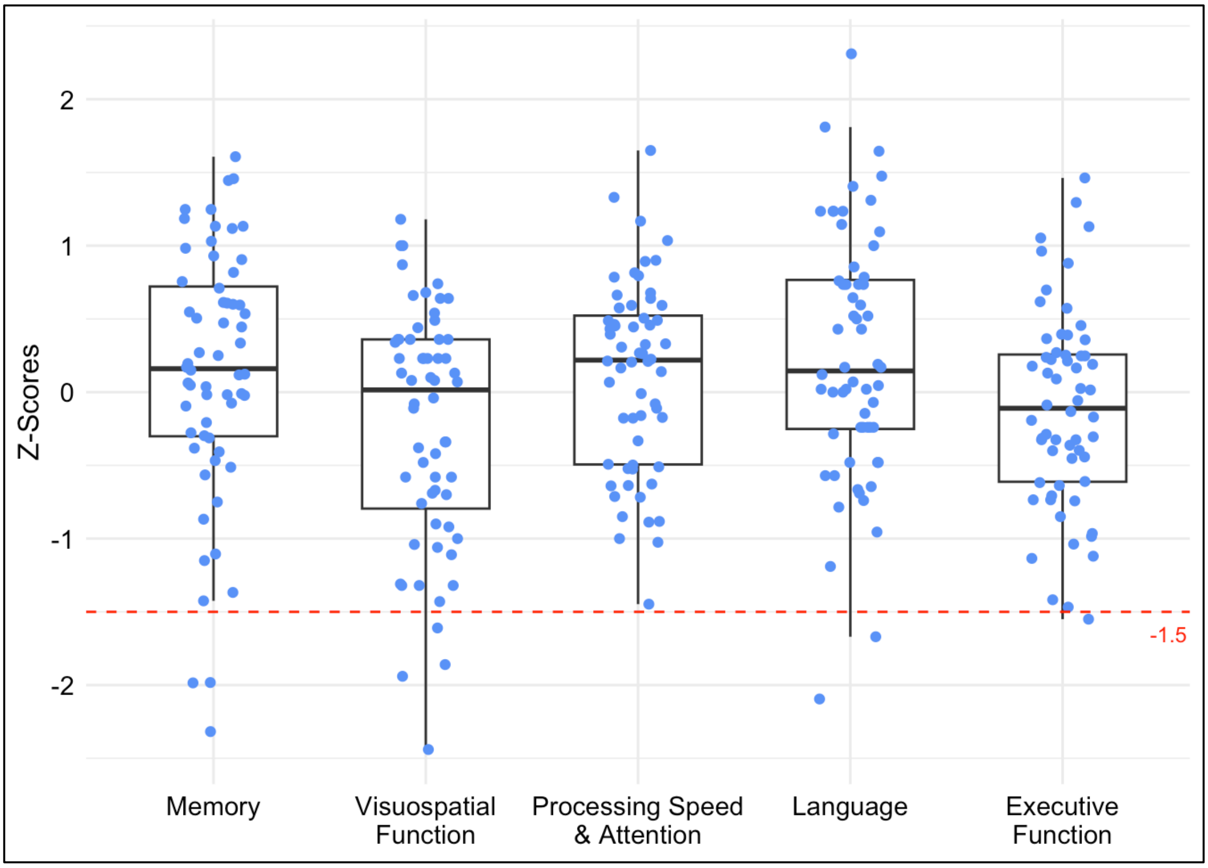


*Note.* The plot marks correspond to individual patients' z-scores averaged across each cognitive domain (*n*=60). Red dotted line corresponds to the psychometric threshold for cognitive impairment (<=-1.5z).

**Table 1**

*Details of normative sample data for psychometric instruments*

| Cognitive domain & test name | Reference | Normative data | Reliability/validity |
|---|---|---|---|
| Processing speed and attention | | | |
| Digit Span forward (DSf) | Wechsler[1] | Wechsler[1] | The subtest has good internal consistency (*r*=.81) and adequate test-retest reliability (*r*=.77). The DS test correlates well with other measures of attention and working memory.[1] |
| Symbol Digit Modalities Test (SDMT) oral version | Smith[2] | Smith[3] | The SDMT correlates well with other measures of processing speed and attention (*r*=.62-.91) and has high test-retest reliability for the oral version (*r*=.76). The oral version correlates well with the written form (*r*>.78). Correlations between the SDMT and the processing speed Wechsler subtests are .62-.91. The task has also been described to measure aspects of attention.[4] |
| Trail Making Test Part A (TMTA) | Reitan[5] | Tombaugh[6] | The TMTA has a high interrater reliability (*r*=.92) and variable test-retest reliability ranging from .46 to .78.[4] |
| Victoria Stroop Test Dots (Stroop D) | Regard[7] | Troyer et al.[8] | The Stroop D measures the speed of naming colours of dots. The Stroop D has high test-retest reliability coefficients (*r*=.90).[4] |

Executive Function

| | | | |
|---|---|---|---|
| Digit Span backward (DSb) | Wechsler[1] | Wechsler[1] | The subtest has good internal consistency (*r*=.82) and adequate test-retest reliability (*r*=.71) The DS test correlates well with other measures of attention and working memory.[1] |
| Letter Fluency Test (OLR) | Delis et al.[9] | Delis et al.[9] | The OLR is traditionally described to examine executive function, involving aspects of language, attention, working memory, cognitive flexibility, and processing speed.[10] The test has high internal consistency and test-retest reliability (*r*=.80-.89). Correlation is high between alternative forms (*r* =.83).[4] |
| Trail Making Test Part B (TMTB) | Reitan[5] | Tombaugh[6] | The TMTB is a measure of mental flexibility and attention, and places demands on executive function. It has high test-retest reliability (*r*=.44-.89), high interrater reliability (*r*=.90) and correlates moderately well with tests of processing speed.[10] |
| Victoria Stroop Test Interference Score (Stroop C/D) | Regard[7] | Troyer et al.[8] | The interference score is traditionally used as a measure of executive function by examining cognitive inhibition.[10] Interference ratio score correlates moderately well with measures of attention ($R^2$=.31) and measure of response inhibition (*r*=.33-.56). Working memory has been described as predictive of Stroop performance. Impaired interference has been described in a multitude of patient groups with known executive dysfunction.[4] |

| | | | |
|---|---|---|---|
| Memory | | | |
| Brief Visuospatial Memory Test-Revised (BVMT-R) | Benedict[11] | Benedict[11] | The BVMT-R measures visuospatial learning and memory, and impaired performance has been reported in patients with memory dysfunction. Test-retest reliability ranges from .60 to .84 across the six available test forms. Alternative forms are thought to be equivalent. The measure correlates with other tests of memory function ($r$=.65-.80).[4] |
| Rey Auditory Verbal Learning Test (RAVLT) | Lezak[12] | Geffen[13] | The RAVLT has good diagnostic utility, correlates moderately well with other measures of verbal memory, and has a high internal reliability ($\alpha$=.90) and adequate test-retest reliability ($r$=.60-.70). Alternative forms are generally reported to have reasonable reliability ($r$>.60). There is a high correlation with total learning score and the delayed score ($r$>.75).[4] |
| Language | | | |
| Category Fluency Test (CFT) | Delis et al.[9] | Delis et al.[9] | The CFT has adequate test-retest reliability ($r$=.70-.79) and marginal internal consistency ($r$=.60-.69). Correlation is moderately high between alternative forms ($r$=.66). There is some association between the CFT and measures of confrontation naming ($r$=.57-.68).[4] |
| Sydney Language Battery Naming Test (SydBat) | Savage et al.[14] | Savage et al.[14] | The SydBat demonstrates good convergent validity, with high correlation between the naming subtest and another visual confrontation naming measure (the Boston Naming Test; $r_s$=.84). The test has high reliability ($\alpha$=.81).[15] |

| | | | |
|---|---|---|---|
| Visuospatial Function | | | |
| Rey-Osterrieth Complex Figure Test (RCFT) Copy | Meyers & Meyers[16] | Mitrushina et al.[10] | The RCFT traditionally examined visuospatial construction. The alternative Taylor figure produces comparable scores.[10] The measure has high interrater reliability (*r*>.90) for total accuracy scores and variable test-retest reliability (*r*=.18-.68). |
| Montreal Cognitive Assessment (MoCA) | Nasreddine et al.[17] | Nasreddine et al.[17] | The MoCA has been validated for an age group of 55-85 years. The test has sensitivity of 90% for detecting mild cognitive impairment and specificity of 87%. The MoCA has demonstrated good test-retest reliability (*r*=.92) and internal consistency ($\alpha$=.83).[17] |
| Test of Premorbid Function (TOPF) | Wechsler[18] | Wechsler[18] | The TOPF is a test of irregular word reading, which is used to estimate premorbid level of function. The test is co-normed with and correlates with WAIS-IV FSIQ (*r*=0.56–.73).[19] |
| SPECTRA: Indices of Psychopathology | Blais and Sinclair[20] | Blais and Sinclair[20] | The SPECTRA is a self-report multiscale measure of adult psychopathology and functioning. All SPECTRA scales demonstrated adequate internal consistency in psychiatric patients ($\alpha$ =.74-.95).[21] SPECTRA scales correlate highly with the PID-5, the PDQ, the SBQ-R, the PAI, the NEO-FFI-3, and the BRIEF-A.[20] |

*Note.* The Digit Span forward and backward is from the Wechsler Adult Intelligence Scale 4th edition (WAIS-IV). The Letter Fluency Test and the Category Fluency Test are from the Delis–Kaplan Executive Function System (D-KEFS) battery. PID-5 = the Personality Inventory for DSM-5; PDQ = the Personality Diagnostic Questionnaire, SBQ-R = the Suicidal Behaviors Questionnaire-Revised; PAI = Personality Assessment Inventory; NEO-FFI-3 = NEO Five Factor Inventory-3; BRIEF-A = Behavior Rating Inventory of Executive Function for Adults

**Table 2**

*Cognitive status classification based on the impression of the treating clinical neuropsychologist*

| Classification category | Description |
|---|---|
| Normal cognition | • No subjective cognitive dysfunction observed by the patient or the carer.<br>• Normal clinical presentation and psychometric performance. |
| Subjective cognitive dysfunction | • Subjective cognitive difficulties observed by the patient or the carer.<br>• Normal clinical presentation and psychometric performance. |
| Mild impairment | • Concern about the patient's cognition from the patient and/or the carer and/or the clinician.<br>• Clinical and/or psychometric evidence of cognitive dysfunction.<br>• Preserved daily function. |
| Moderate impairment | • Concern about the patient's cognition from the patient and/or the carer and/or the clinician.<br>• Clinical and/or psychometric evidence of cognitive impairment beyond secondary dysfunction and consistent with a pattern of primary impairment.<br>• Evidence of functional impairment. |

*Note.* Subjective cognitive difficulties refer to a patient's or a carer's perceived cognitive change. Criteria for mild and moderate impairment was partially adopted from the DSM-V neurocognitive disorder criteria.[22]

**Table 3**

*Therapy characteristics*

| Types of prior therapy | | | | | |
|---|---|---|---|---|---|
| Therapy | *N* | % | Therapy | *N* | % |
| Chemotherapy | 60 | 100.00 | CNS-penetrating radiotherapy | 6 | 10.00 |
| Corticosteroids | 57 | 95.00 | Antibody-drug conjugates | 4 | 6.67 |
| Monoclonal antibodies | 54 | 90.00 | AlloSCT | 4 | 6.67 |
| Radiotherapy | 33 | 55.00 | Immunomodulatory drugs | 3 | 5.00 |
| CNS-penetrating chemotherapy | 39 | 65.00 | CNS-directed radiotherapy | 2 | 3.33 |
| AutoSCT | 17 | 28.33 | Proteasome inhibitor | 2 | 3.33 |
| Bispecific antibodies | 11 | 18.33 | CAR-T | 2 | 3.33 |
| BTK-inhibitors | 8 | 13.33 | Pl3-kinase inhibitor | 1 | 1.67 |
| BCL2 family inhibitors | 6 | 10.00 | Checkpoint inhibitors | 1 | 1.67 |
| **Types of bridging therapy (n = 51)** | | | | | |
| Therapy | *N* | % | Therapy | *N* | % |
| Radiotherapy | 11 | 21.57 | BTK-inhibitor | 3 | 5.88 |
| Chemotherapy | 9 | 17.65 | Antibody-drug conjugates | 2 | 3.92 |
| Chemotherapy + radiotherapy | 7 | 13.73 | Monoclonal antibodies + immunomodulating agents + corticosteroids | 2 | 3.92 |
| Corticosteroids | 5 | 9.80 | Monoclonal antibodies | 1 | 1.96 |
| Radiotherapy + corticosteroids | 5 | 9.80 | Chemotherapy + BTK-inhibitor | 1 | 1.96 |
| Chemotherapy + corticosteroids | 4 | 7.84 | Radiotherapy + BTK-inhibitor | 1 | 1.96 |
| **Pre-CART response to therapy** | | | | | |
| Response type | *N* | % | Response type | *N* | % |
| Progressive disease | 30 | 50.00 | Complete response | 10 | 16.67 |
| Partial response | 13 | 21.67 | Stable disease | 7 | 11.67 |
| **Planned CAR-T product** | | | | | |
| Product | *N* | % | Product | *N* | % |
| SOC axicabtagene ciloleucel | 30 | 50.00 | SOC brexucabtagene autoleucel | 3 | 5.00 |

| | | | | | |
|---|---|---|---|---|---|
| SOC tisagenlecleucel | 15 | 25.00 | Clinical trial products | 12 | 20.00 |

*Note.* tf = transformed disease; CNS = central nervous system; AutoSCT = autologous stem cell transplantation; AlloSCT = allogeneic stem cell transplantation; BTK = Bruton's tyrosine kinase; BCL2 = B-cell lymphoma 2; CAR-T = chimeric antigen receptor T-cell therapy; Pl3 = phosphoinositide 3-kinase; SOC = standard of care.

**Supplementary references**


1. Wechsler D. Wechsler Adult Intelligence Scale: Technical and interpretive manual. 4th ed. San Antonio: Pearson Assessment; 2008.
2. Smith A. Symbol digit modalities test. Los Angeles: Western psychological services; 1973. 22 p.
3. Smith A. Symbol digits modalities test: Manual (10th printing). Western Psychological Services: Los Angeles. 2007.
4. Strauss E, Sherman EM, Spreen OA. Compendium of neuropsychological tests: Administration, norms, and commentary. American chemical society; 2006.
5. Reitan RM. The Relation of the Trail Making Test to organic brain damage. J Consult Psychol. 1955;19(5):393–4.
6. Tombaugh TN. Trail Making Test A and B: Normative data stratified by age and education. Arch Clin Neuropsychol [Internet]. 2004 Mar 1 [cited 2023 May 30];19(2):203–14. Available from: https://www.sciencedirect.com/science/article/pii/S0887617703000398
7. Regard M. Cognitive rigidity and flexibility: A neuropsychological study. University of Victoria. Victoria. 1981.
8. Troyer AK, Leach L, Strauss E. Aging and response inhibition: Normative data for the Victoria Stroop Test. Neuropsychol Dev Cogn B Aging Neuropsychol Cogn [Internet]. 2006 Jan 1 [cited 2023 May 30];13(1):20–35. Available from: https://doi.org/10.1080/138255890968187
9. Delis DC, Kaplan E, Kramer JH. Delis-Kaplan executive function system. Assessment. 2001.
10. Mitrushina M, Boone KB, Razani J, D'Elia LF. Handbook of Normative Data for Neuropsychological Assessment. Oxford University Press; 2005. 1052 p.
11. Benedict RH. Brief visuospatial memory test--revised. PAR; 1997.
12. Lezak MD. Neuropsychological assessment. 2nd ed. New York: Oxford University Press; 1983.
13. Geffen G, Moar KJ, O'hanlon AP, Clark CR, Geffen LB. Performance measures of 16– to 86-year-old males and females on the auditory verbal learning test. Clin Neuropsychol [Internet]. 1990 Mar 1 [cited 2023 May 30];4(1):45–63. Available from: https://doi.org/10.1080/13854049008401496
14. Savage S, Hsieh S, Leslie F, Foxe D, Piguet O, Hodges JR. Distinguishing subtypes in primary progressive aphasia: application of the Sydney Language Battery. Dement Geriatr Cogn Disord [Internet]. 2013 [cited 2023 May 30];35(3–4):208–18. Available

from: https://search.ebscohost.com/login.aspx?direct=true&AuthType=sso&db=mnh&AN=23467307&site=ehost-live&custid=s2775460

15. Janssen N, Roelofs A, van den Berg E, Eikelboom WS, Holleman MA, in de Braek DMJM, et al. The diagnostic value of language screening in primary progressive aphasia: Validation and application of the Sydney Language Battery. J Speech Lang Hear Res [Internet]. 2022 Jan [cited 2023 May 30];65(1):200–14. Available from: https://search.ebscohost.com/login.aspx?direct=true&AuthType=sso&db=a2h&AN=154696192&site=ehost-live&custid=s2775460
16. Meyers JE, Meyers KR. Rey complex figure test under four different administration procedures. Clin Neuropsychol [Internet]. 1995 Feb 1 [cited 2023 May 30];9(1):63–7. Available from: https://doi.org/10.1080/13854049508402059
17. Nasreddine ZS, Phillips NA, Bédirian V, Charbonneau S, Whitehead V, Collin I, et al. The Montreal Cognitive Assessment, MoCA: A Brief Screening Tool For Mild Cognitive Impairment. Journal of the American Geriatrics Society [Internet]. 2005 [cited 2023 Mar 31];53(4):695–9. Available from: https://onlinelibrary.wiley.com/doi/abs/10.1111/j.1532-5415.2005.53221.x
18. Wechsler D. Advanced clinical solutions for the WAIS-IV and WMS-IV. San Antonio: The Psychological Corporation. 2009.
19. Shura RD, Ord AS, Martindale SL, Miskey HM, Taber KH. Test of Premorbid Functioning: You're doing it wrong, but does it matter? Arch Clin Neuropsychol [Internet]. 2022 Jul 19 [cited 2023 Mar 31];37(5):1035–40. Available from: https://academic.oup.com/acn/article/37/5/1035/5836861
20. Blais MA, Sinclair SJ. SPECTRA Indices of Psychopathology. PAR; 2018.
21. Blais MA, Sinclair SJ, Richardson LA, Massey C, Stein MB. External correlates of the SPECTRA: Indices of psychopathology (SPECTRA) in a clinical sample. Clin Psychol Psychother [Internet]. 2021 [cited 2023 May 30];28(4):929–38. Available from: https://onlinelibrary.wiley.com/doi/abs/10.1002/cpp.2546
22. American Psychiatric Association, American Psychiatric Association, editors. Diagnostic and statistical manual of mental disorders: DSM-5. 5th ed. Arlington, VA: American Psychiatric Association; 2013. 947 p.
